# Evaluating Large Language Models in Extracting Cognitive Exam Dates and Scores

**DOI:** 10.1101/2023.07.10.23292373

**Authors:** Hao Zhang, Neil Jethani, Simon Jones, Nicholas Genes, Vincent J. Major, Ian S. Jaffe, Anthony B. Cardillo, Noah Heilenbach, Nadia Fazal Ali, Luke J. Bonanni, Andrew J. Clayburn, Zain Khera, Erica C. Sadler, Jaideep Prasad, Jamie Schlacter, Kevin Liu, Benjamin Silva, Sophie Montgomery, Eric J. Kim, Jacob Lester, Theodore M. Hill, Alba Avoricani, Ethan Chervonski, James Davydov, William Small, Eesha Chakravartty, Himanshu Grover, John A. Dodson, Abraham A. Brody, Yindalon Aphinyanaphongs, Arjun Masurkar, Narges Razavian

## Abstract

**Importance:** Large language models (LLMs) are crucial for medical tasks. Ensuring their reliability is vital to avoid false results. Our study assesses two state-of-the-art LLMs (ChatGPT and LlaMA-2) for extracting clinical information, focusing on cognitive tests like MMSE and CDR.

**Objective:** Evaluate ChatGPT and LlaMA-2 performance in extracting MMSE and CDR scores, including their associated dates.

**Methods:** Our data consisted of 135,307 clinical notes (Jan 12th, 2010 to May 24th, 2023) mentioning MMSE, CDR, or MoCA. After applying inclusion criteria 34,465 notes remained, of which 765 underwent ChatGPT (GPT-4) and LlaMA-2, and 22 experts reviewed the responses. ChatGPT successfully extracted MMSE and CDR instances with dates from 742 notes. We used 20 notes for fine-tuning and training the reviewers. The remaining 722 were assigned to reviewers, with 309 each assigned to two reviewers simultaneously. Inter-rater-agreement (Fleiss’ Kappa), precision, recall, true/false negative rates, and accuracy were calculated. Our study follows TRIPOD reporting guidelines for model validation.

**Results:** For MMSE information extraction, ChatGPT (vs. LlaMA-2) achieved accuracy of 83% (vs. 66.4%), sensitivity of 89.7% (vs. 69.9%), true-negative rates of 96% (vs 60.0%), and precision of 82.7% (vs 62.2%). For CDR the results were lower overall, with accuracy of 87.1% (vs. 74.5%), sensitivity of 84.3% (vs. 39.7%), true-negative rates of 99.8% (98.4%), and precision of 48.3% (vs. 16.1%). We qualitatively evaluated the MMSE errors of ChatGPT and LlaMA-2 on double-reviewed notes. LlaMA-2 errors included 27 cases of total hallucination, 19 cases of reporting other scores instead of MMSE, 25 missed scores, and 23 cases of reporting only the wrong date. In comparison, ChatGPT’s errors included only 3 cases of total hallucination, 17 cases of wrong test reported instead of MMSE, and 19 cases of reporting a wrong date.

**Conclusions:** In this diagnostic/prognostic study of ChatGPT and LlaMA-2 for extracting cognitive exam dates and scores from clinical notes, ChatGPT exhibited high accuracy, with better performance compared to LlaMA-2. The use of LLMs could benefit dementia research and clinical care, by identifying eligible patients for treatments initialization or clinical trial enrollments. Rigorous evaluation of LLMs is crucial to understanding their capabilities and limitations.

## Introduction

Large-scale language models (LLMs) [1–4] have emerged as powerful tools in natural language processing (NLP), capable of performing diverse tasks when prompted [5] [6]. These models have demonstrated impressive clinical reasoning abilities [7], successfully passing medical licensing exams [8] [9] [10] and generating medical advice on distinct subjects, including cardiovascular disease [11], breast cancer [12], colonoscopy [13], and general health inquiries [14], [6], [15] [16]. These models can produce clinical notes [16] and assist in writing research articles [16]. Medical journals have begun developing policies around use of LLMs in writing [17] [18] [19] [20] [21] [22] and reviewing. Examples of such LLMs include ChatGPT [2] [1], Med-PALM-2 [3], LlaMA-2 [4], and open-source models actively produced by the community [23].

In this study, we focus on evaluating *information extraction* abilities of Large Language Models from clinical notes, specifically focusing on proprietary ChatGPT (powered by GPT-4 [2]), and open source LlaMA-2 [4] LLMs. Information extraction involves the retrieval of specific bits of information from unstructured clinical notes, a task historically handled by rule-based systems [24,25] [26] [27] [28] [29] [30] or language models explicitly trained on datasets annotated by human experts [31] [32] [33] [34] [35] [36]. Rule-based systems lack a contextual understanding and struggle with complex sentence structures, ambiguous language, and long-distance dependencies, often leading to high false positive rates and low sensitivities [37] [38] [39] [40]. Additionally, training a new model for this task can be computationally demanding and require substantial human effort. In contrast, LLMs, such as ChatGPT or LlaMA-2, operate at “zero-shot” capacity [41] [42] [43], i.e., only requiring a prompt describing the desired information to be extracted.

Despite their promise, LLMs also have a potential limitation - the generation of factually incorrect yet highly convincing outputs, commonly known as “hallucination.” The massive architectures and complex training schemes of LLMs hamper “model explanation” and the ability to intrinsically guarantee behavior. This issue has been extensively discussed in the literature, emphasizing the need for cautious interpretation and validation of information generated by LLMs [44] [2] [45].

One area where LLMs may greatly benefit healthcare is in the identification of memory problems and other symptoms indicative of Alzheimer’s Disease and Alzheimer’s Disease Related Dementias (AD/ADRD) within clinical notes. AD/ADRD is commonly underdiagnosed or diagnosed later in the disease trajectory, particularly in racial and ethnic minoritized groups [46] [47] [48] [49] [50] [51]. The precise extraction of cognitive test scores holds significant importance in the development and clinical validation of tools that can facilitate early detection [52] of AD/ADRD in the clinic. Earlier identification can lead to a host of benefits, including assisting with advanced care planning, performing secondary cardiovascular disease prevention, which may reduce worsening of cognitive impairment [53] [54], identification for serving in research trials [55] [56,57], and with the rapid advancement in biologic therapeutics, the opportunity to receive potentially disease modifying drugs [57] [58]. Accurately extracting cognitive exam scores (often buried in clinical notes and not documented in any structured field), enables validation, training and fine-tuning of models at a much larger scale in a clinical setting for a much more racial/ethnically diverse patient population set compared to current research cohorts.

The primary focus of this paper is therefore on the validation of two state-of-the-art LLMs (ChatGPT powered by GPT-4, and LlaMA-2), for information extraction related to cognitive tests, specifically the Mini-Mental State Examination (MMSE) [59] and Clinical Dementia Rating (CDR) [60], from clinical notes of a racially and ethnically diverse patient population. Our objective is to accurately extract all instances of (the exam score, and the date when the exam was administered) using these LLMs.

This study represents a large-scale formal evaluation of two state of the art LLMs (ChatGPT, and LlaMA-2) performance in information extraction from clinical notes. Going forward, we intend to employ this benchmark dataset to validate other (open or closed-source) LLMs. Furthermore, we plan to adopt a similar approach to validate LLMs for information extraction across various clinical use cases. By prioritizing prompt engineering with ChatGPT and LlaMA-2 for extracting clinical information, this research aims to enhance our understanding of the potential of LLMs in healthcare and facilitate the development of reliable and robust clinical information extraction tools.

## Methods

This study is approved under IRB i20-01095, “Understanding and predicting Alzheimer’s Disease.” NYU DataCore services were utilized to prepare the data as described below. A HIPAA-compliant private instance of ChatGPT was utilized for this study. LlaMA-2 (“Llama-2-70b-chat” version) was evaluated on two A100 Nvidia GPUs on our local high performance computing servers. This Diagnostic/Prognostic study designed to validate the diagnostic accuracy of two LLMs (ChatGPT and LlaMA-2) in extracting cognitive exam dates and scores, follows the follows the TRIPOD Prediction Model Validation reporting guidelines.

### Dataset

An original cohort of 135,307 clinical notes corresponding to inpatient, outpatient, and emergency department visits between January 12th 2010 and May 24th 2023, which included any of the following keywords (‘MMSE’, ‘CDR,’ or ‘MoCA’ case-insensitive) were identified (see Figure 1). MMSE stands for Mini Mental State Exam, CDR stands for Cognitive Dementia Rating, and MoCa stands for Montreal Cognitive Assessment [61]. These notes belonged to 52,948 patients. From among these patients, 26,355 had a non-contrast brain Magnetic Resonance Imaging (MRI) in the system. Limiting the clinical notes to those who had an MRI in the system resulted in 77,547 notes. These notes were extracted. At this stage we further limited the notes to those including any mentions of MMSE and/or CDR (ignoring MoCA), which yielded 34,465 clinical notes for analysis.

**Figure 1:**
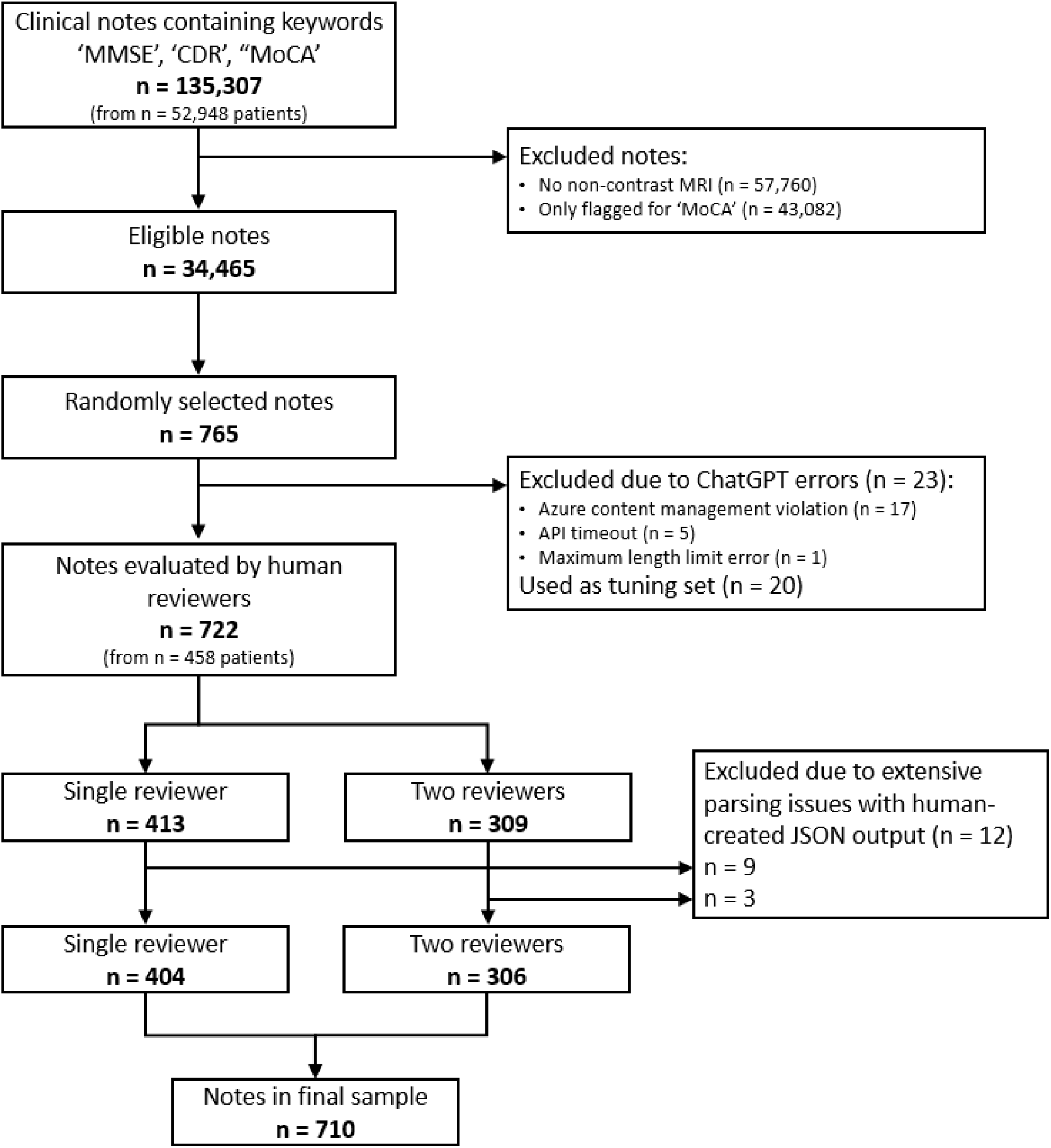
Flowchart of clinical notes evaluated for inclusion in the final sample of GPT-analyzed notes.

The choice for requiring patients to have a brain MRI as well as MMSE and/or CDR enables us to have a similar level of granularity as the Alzheimer’s Disease Neuro-Imaging Initiative (ADNI) [62], which also uses MMSE and CDR for definition of mild cognitive impairment and dementia stages. This further enables us to harmonize our clinical dataset with these large research cohorts. To elucidate the impact of this choice (restriction of cohort to those with MRI) on the racial breakdown of our study, we include a demographics comparison between the two sets (original 52,948 patients, and the 26,355 with an MRI) in the supplementary section S1.

Similarly, the choice to ignore MoCA was due to the lack of inclusion of MoCA in standard definition for stages of cognitive impairment in ADNI. The mild cognitive impairment and (mild, moderate or severe) dementia definition criteria utilized in ADNI are included in Supplementary Table S1. Data harmonization is beyond the scope of this paper, although information extraction plays a substantial role in enabling it.

From among 34,465 notes that fit the inclusion criteria, a random selection of 765 notes was identified to undergo information extraction via ChatGPT and manual evaluation. 765 was the total number of the notes needed to satisfy two conditions: 1) Each reviewer not being assigned more than 50 notes to review, and 2) at least around 15 notes per reviewer being double-reviewed by another random reviewer. From among these 765 notes, ChatGPT encountered application programming interface (API) errors in 23 cases (3%). These errors arose from “Azure content management violations’’ [63] (17 cases), API timeouts (5 cases), and maximum length limit errors (1 case). Supplementary Table S2 includes a more detailed description of these errors. The remaining 742 were considered for assignment to domain expert reviewers, and underwent analysis by LlaMA-2.

### Generative AI, ChatGPT

ChatGPT (GPT-4, API version “2023-03-15-preview”) was used on these 765 notes to extract all instances of the cognitive tests—MMSE and CDR—along with the dates at which the tests were mentioned to have been administered. Two examples of our task are provided in the supplementary section S2. Inference was successful for 742 notes. The complete API call, along with the exact prompt, the temperature, and other hyper-parameters are included in Supplementary Table S3. The prompt included a request to return these results in a JSON format. ChatGPT’s response (full), as well as the JSON formatted dialogue response were recorded in one session on June 9th 2023. The notes sent to ChatGPT were text-only, stripped of the rich-text formatting (RTF) native to our EHR system (Epic Systems, Verona, WI). This reduced token count by approximately ten-fold, enabling notes to fit into the GPT4-8K input window and removing a substantial source of confusion for the LLM in prompt tuning. The date that the encounter was recorded in Epic was appended at the beginning of the note, proceeding with a column (“:”) then the note text. See Supplementary Table S3 for the API request, including the prompt.

### Generative AI, LlaMA-2

We used LlaMA-2 (version “Llama-2-70b-chat”) on all the notes which ChatGPT produced valid answer. All pre-processing steps on the notes were similar to that of ChatGPT. The context window was limited to the first 3696 tokens. The complete API call, along with the exact prompt, the temperature, and other hyper-parameters are included in Supplementary Table S4.

### Hyper-parameter and Prompt Tuning

For both ChatGPT and LlaMA-2, we assigned 20 notes out of the 742 as our hyper-parameter and prompt tuning set. For ChatGPT, an interactive cloud-based environment (i.e playground) was utilized initially to fine-tune the prompt. After initial exploratory analysis using these 20 notes, they were scored via the API using the best prompt and hyper-parameter found in the interactive mode. For LlaMA-2, the exploration was performed locally, on the same 20 notes. All human expert reviewers (detailed below) were instructed to first review the ChatGPT results of the 20 cases in a RedCap survey. The goal of this step was to train the reviewers, refine the information presented in RedCap, improve clarification of the questions, and potentially refine the prompt. These 20 notes were then excluded from any additional analysis.

### Human Expert Reviewers

Our team included 22 medically trained expert reviewers who volunteered and were trained to review an (HTML formatted) note, provide ground truth, and judge the correctness and completeness of ChatGPT answers for each cognitive test. Fully (HTML) formatted notes were pulled using an Epic web service, and were fed into the RedCap survey. Redcap survey rendered the note’s HTML formatting, to ensure notes could be displayed to users in the same format as the readers are accustomed to seeing them clinically, rather than the text-only, computer-friendly format provided to GPT. For 21 of these reviewers, each reviewer was assigned approximately 50 clinical notes to evaluate. From among each reviewer’s 50 assigned notes, about 15 notes were assigned to another random reviewer. The assignment algorithm randomly selected a pair of reviewers for each of our 309 double-reviewed notes and assigned the remaining notes to a randomly selected reviewer until each reviewer reached 50 notes or we fully assigned all notes. This random assignment was a necessary step for ensuring correctness of Fleiss’ Kappa [64] metric for inter-rater-agreement. As a result, there was a slight variation in the total number of assigned notes for each reviewer.

Overall, 722 notes were assigned to these 21 reviewers, of which 309 were double-reviewed and 413 were solo-reviewed. The double-reviewed 309 notes were utilized in reporting inter-rater-agreement metrics. After the review, 69 out of 309 notes had at least one disagreement between the two reviewers based on one of the four questions: *Whether ChatGPT’s response on MMSE was correct; whether ChatGPT’s response on MMSE included all instances of MMSE found in the clinical note; whether ChatGPT’s response on CDR was correct; and whether ChatGPT’s response on CDR included all instances of CDR found in the clinical note*. A 22nd reviewer was then tasked to review these 69 notes again to provide a third review. Majority vote was then employed to identify the final answer and the ground truth provided by the reviewer whose answer was in the majority vote was used to calculate detailed precision/recall metrics. When both reviewers fully agreed and their JSON results were both valid for analysis, we randomly selected one to compute the precision and recall. Details of the parsing of the JSON result are included in the supplementary section S3. These expert-provided ground truth results were the basis for evaluating LlaMA-2.

### Statistical Approach

For double-reviewed notes, we reported Fleiss Kappa [64] as a measure for inter-rater-agreement, for ChatGPT analysis. We reported this metric for the four questions (Is MMSE complete/correct, and is CDR complete/correct). Additionally, for double-reviewed notes, we computed a 2-way Fleiss Kappa for MMSE and CDR lists of (outcome and date) tuples extracted from the JSON responses of expert reviewers, comparing them against each other, to derive inter-rater-agreement. Fleiss’ Kappa is useful when the assignment of a note to reviewer pairs has been random (uniform), and each note has been reviewed by a subset of reviewers [65] [66]. We only considered exact matches (i.e [MMSE-27/30, date “10-10-2010”], with [MMSE-26/30, date “10-10-2010”] is just as bad as [MMSE-5/30, date “10-10-2012”]). Kappa can be interpreted as follows: 40%–59% would be *Weak*, 60%–79% would be *Moderate*, 80%– 90% would be Strong, and Above 90% would be *Almost Perfect [65]*. In addition to 2-way Kappa, we also report a 3-way Kappa on the entries of MMSE and CDR results extracted from the JSON results, computing the joint agreement between the results of ChatGPT and the results provided by two human reviewers.

We also report per test type (MMSE and CDR), Accuracy, True and False Negative Rates, Micro- and Macro-Precision and Micro- and Macro-Recall for both ChatGPT and LlaMA-2. Accuracy is defined as the percentage of correct results (at clinical note level), correct being defined as the list of (Value/Date) tuples in the JSON entries for the LLM and Ground Truth being fully identical. Macro-Precision for MMSE (or CDR) is the average (at the note level) of percentage of correct MMSE (or CDR) tuples extracted (correct both in date and score values compared to an entry mentioned in the ground truth for MMSE (or CDR)). Macro-Recall for MMSE (or CDR) is the average (at the note level) of the percentage of the MMSE items in the ground truth that are extracted by the LLM. *Micro-*precision is calculated as percentage of *correct* MMSE (or CDR) items extracted by the LLM, from among all extracted MMSE (or CDR) items by that LLM, and is calculated as one number across all notes combining all notes’ entries. Micro-recall is similarly calculated as the percentage of all MMSE (or CDR) items mentioned in the ground truth that were extracted by the LLM.

## Results

ChatGPT analyzed 765 notes for extraction of Mini Mental Status Exam (MMSE) and Cognitive Dementia Rating (CDR) scores and exam dates. Of these, 23 encountered API error (3%), and 20 were used to fine-tune prompt and hyper-parameters. The remaining 722 notes were assigned to human expert reviewers who manually reviewed (and provided ground truth for) these notes. LlaMA-2 analyzed these 722 notes as well. Characteristics of these 722 notes and associated patients are included in Table 1.

**Table 1:**
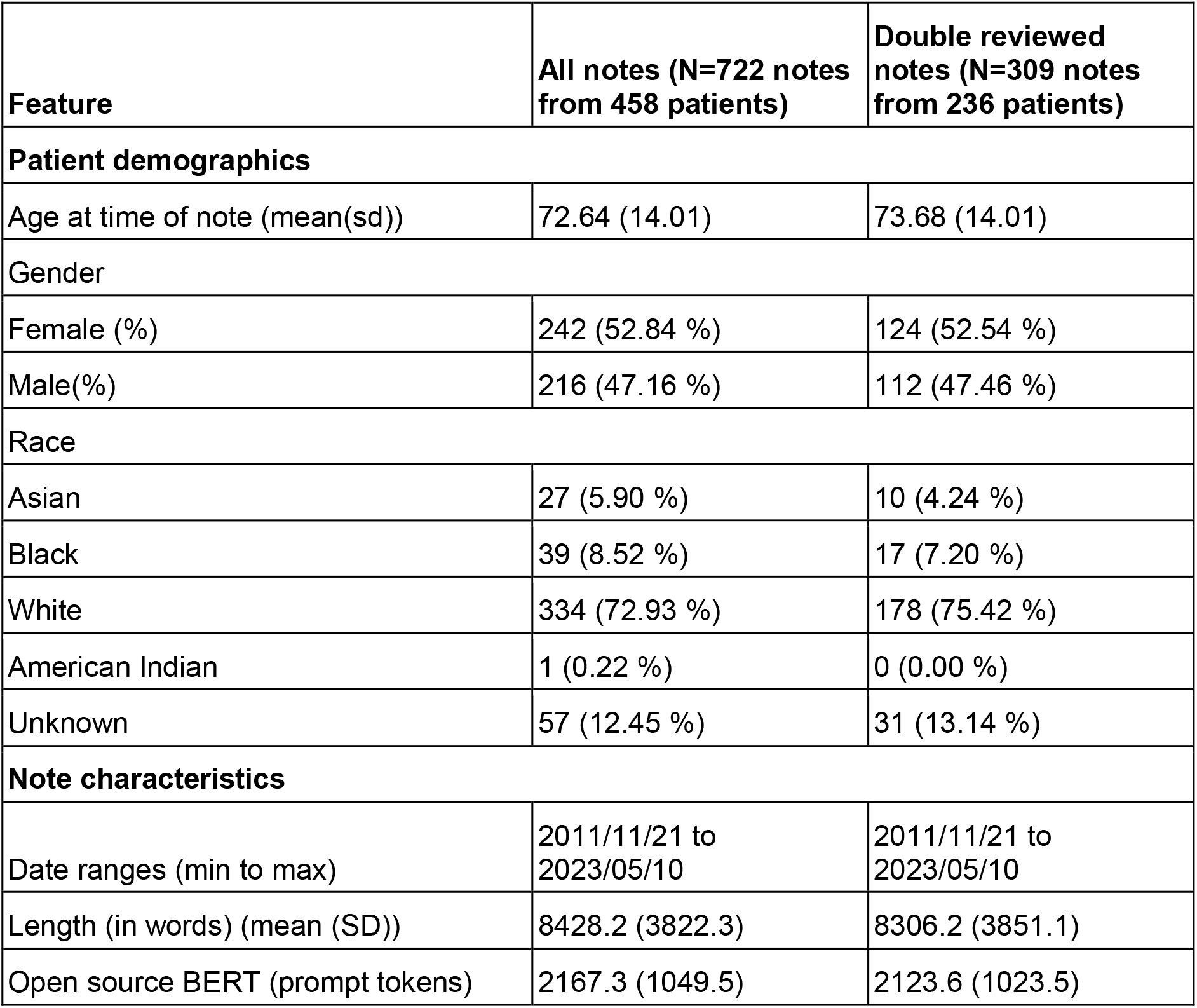
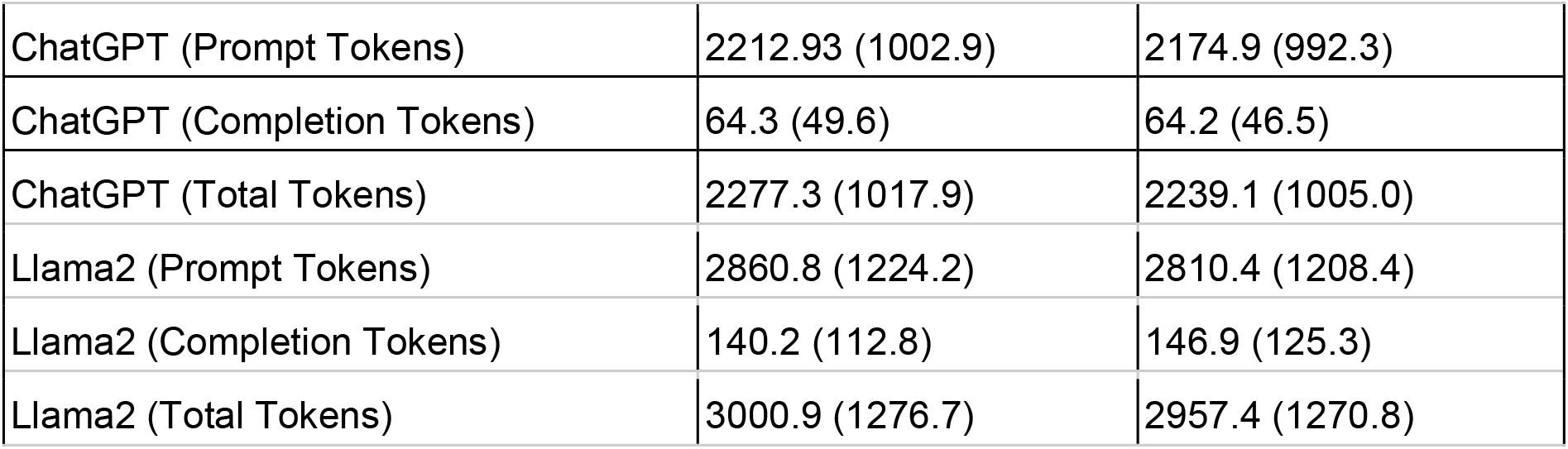
Characteristics of 722 notes which are manually evaluated, and their corresponding patients.

| Feature | All notes (N=722 notes from 458 patients) | Double reviewed notes (N=309 notes from 236 patients) |
| --- | --- | --- |
| **Patient demographics** |  |  |
| Age at time of note (mean(sd)) | 72.64 (14.01) | 73.68 (14.01) |
| Gender |  |  |
| Female (%) | 242 (52.84 %) | 124 (52.54 %) |
| Male(%) | 216 (47.16 %) | 112 (47.46 %) |
| Race |  |  |
| Asian | 27 (5.90 %) | 10 (4.24 %) |
| Black | 39 (8.52 %) | 17 (7.20 %) |
| White | 334 (72.93 %) | 178 (75.42 %) |
| American Indian | 1 (0.22 %) | 0 (0.00 %) |
| Unknown | 57 (12.45 %) | 31 (13.14 %) |
| **Note characteristics** |  |  |
| Date ranges (min to max) | 2011/11/21 to 2023/05/10 | 2011/11/21 to 2023/05/10 |
| Length (in words) (mean (SD)) | 8428.2 (3822.3) | 8306.2 (3851.1) |
| Open source BERT (prompt tokens) | 2167.3 (1049.5) | 2123.6 (1023.5) |
| ChatGPT (Prompt Tokens) | 2212.93 (1002.9) | 2174.9 (992.3) |
| ChatGPT (Completion Tokens) | 64.3 (49.6) | 64.2 (46.5) |
| ChatGPT (Total Tokens) | 2277.3 (1017.9) | 2239.1 (1005.0) |
| Llama2 (Prompt Tokens) | 2860.8 (1224.2) | 2810.4 (1208.4) |
| Llama2 (Completion Tokens) | 140.2 (112.8) | 146.9 (125.3) |
| Llama2 (Total Tokens) | 3000.9 (1276.7) | 2957.4 (1270.8) |

Of the double-reviewed 309 notes, 69 had at least one disagreement between the responses to the four questions (if ChatGPT’s response for MMSE/CDR is correct/complete) and were assigned to a new reviewer for a third opinion. Among the responses with disagreement, 9 disagreed about correctness of MMSE answers, 40 disagreed about completeness of MMSE answers, 17 disagreed about correctness of CDR answers, and 22 disagreed about completeness of CDR answers. The average response (at the note level) by the included reviews for the four yes/no questions are included in Table 2. Overall reviewers considered ChatGPT’s response to be 96.5% and 98% correct for MMSE and CDR respectively. The assessment for whether ChatGPT’s answers are also complete (i.e. they do not miss anything) was slightly lower averaging about 84% and 83% for MMSE and CDR respectively.

**Table 2:** Average response (at the note level) of the responses of reviewers in judging if ChatGPT’s answers for MMSE and CDR are correct and/or complete.

|  | All notes (N=722) | Double reviewed notes (N=309) |
| --- | --- | --- |
| Is ChatGPT's answer for MMSE correct? (%) | 96.5 (sd 18.2) | 96.4 (sd 18.5) |
| Is ChatGPT's answer for MMSE complete? (%) | 85.0 (sd 35.7) | 84.7 (sd 36.0) |
| Is ChatGPT's answer for CDR correct? (%) | 98.0 (sd 13.7) | 99.6 (sd 5.6) |
| Is ChatGPT's answer for CDR complete? (%) | 80.4 (sd 39.6) | 83.4 (sd 37.1) |

The inter-rater-agreements between reviewers were calculated based on Fleiss’ Kappa and are summarized in Table 3. In addition to measuring Fleiss’ Kappa between reviewers based on double-reviewed notes (reported as 2-way Fleiss’ Kappa in Table 3), we also report agreement between ChatGPT, and the two human reviewers (reported as 3-way Fleiss’ Kappa in Table 3). The 2-way agreement on the yes/no questions was high (94% agreement between reviewers for MMSE and 89% agreement for CDR). There was some disagreement in judging the completeness of the answer, leading to a Kappa value of 75% for MMSE (and 85% for CDR). More notably, when analyzing the elements of the ground truth JSON, the 2-way agreement was excellent both for scores (83% for MMSE and 80% for CDR) and for dates (93% for MMSE and 79% for CDR). When measuring the 3-way agreement, there was an increase in all the metrics except MMSE dates. The accuracy and results of JSON formatting of the responses are included in supplementary section S4.

**Table 3:**
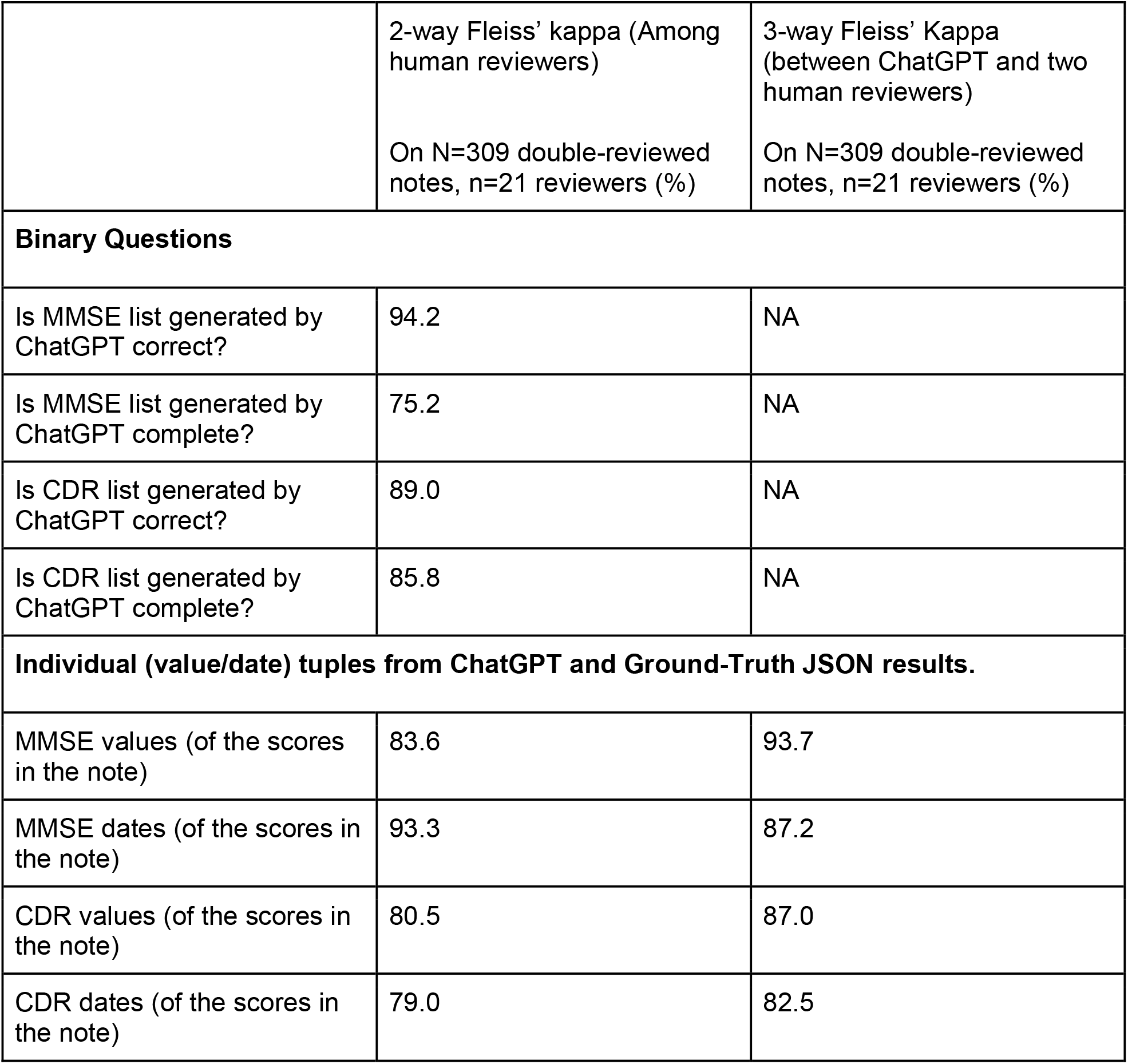
Fleiss’ kappa inter-rater-agreement metric between reviewers (2-way) and reviewers and ChatGPT (3-way) over the double-reviewed notes.

**Table 4:** Aggregate Accuracy, True Negative Rate, (Micro- and Macro-) Precision and Recall for MMSE and CDR scores extracted by ChatGPT and LlaMA-2.

|  | All notes with parsed JSON (N=710) |  | Double-reviewed notes with parsed JSON (N=306) |  |
| --- | --- | --- | --- | --- |
|  | ChatGPT | LLaMA-2 | ChatGPT | LLaMA-2 |
| **MMSE** |  |  |  |  |
| Total notes without any MMSE (in ground truth) | 115 |  | 48 |  |
| Total notes without any MMSE (in GPT results) | 77 | 110 | 25 | 46 |
| Total correctly predicted empty MMSEs | 76 | 66 | 24 | 23 |
| ChatGPT's True Negative Rate for MMSE(%) | 98.7 | 60.0 | 96 | 50.0 |
| ChatGPT's False Negative Rate for MMSE(%) | 1.2 | 40.0 | 4 | 50.0 |
| Remaining notes with un-empty GPT response undergone Precision/Recall calculation for MMSE | 633 | 600 | 281 | 260 |
| Total MMSE instances predicted | 831 | 957 | 366 | 410 |
| MMSE Macro Precision (mean % (sd %)) | 82.9 (sd 36.2) | 62.2(sd 45.5) | 82.7 (sd 36.8) | 63.4 (sd 44.9) |
| MMSE Macro Recall (mean % (sd %)) | 87.8 (sd 30.4) | 69.9 (sd 43.5) | 89.7 (sd 28.3) | 71.8 (sd 42.1) |
| MMSE Micro Precision (%) | 83.8 | 57.7 | 84.1 | 59.3 |
| MMSE Micro Recall (%) | 83.7 | 68.1 | 87.5 | 69.0 |
| Total notes with any error MMSE result | 121 | 238 | 52 | 98 |
| Overall accuracy of MMSE (%) | 82.9 | 66.4 | 83.0 | 68.0 |
| **CDR** |  |  |  |  |
| Total notes without CDR (in ground truth) | 608 |  | 260 |  |
| Total notes without CDR (in GPT results) | 533 | 497 | 233 | 215 |
| Total correctly predicted empty CDR | 532 | 489 | 233 | 212 |
| CDR True Negative Rate (%) | 99.8 | 98.4 | 100 | 98.6 |
| CDR False Negative Rate (%) | 0.2 | 1.6 | 0 | 1.4 |
| Remaining notes with un-empty GPT response undergone Precision/Recall calculation for CDR | 177 | 213 | 73 | 153 |
| Total CDR instances predicted | 256 | 344 | 92 | 153 |
| CDR Macro Precision (mean % sd %) | 48.3 (sd 49.9) | 16.1 (sd 35.5) | 57.5 (sd 49.4) | 18.1 (sd 36.9) |
| CDR Macro Recall (mean % sd %) | 84.3 (sd 36.3) | 39.7 (sd 48.7) | 91.3 (sd 28.1) | 43.5 (sd 49.6) |
| CDR Micro Precision (%) | 36.3 | 12.0 | 51.0 | 13.2 |
| CDR Micro Recall (%) | 85.3 | 37.6 | 92.1 | 39.2 |
| Total notes with any error CDR result | 91 | 181 | 31 | 76 |
| Overall accuracy of CDR (%) | 87.1 | 74.5 | 89.8 | 75.4 |

ChatGPT had an excellent True Negative Rate—over 96% for MMSE and 100% for CDR in double-reviewed notes. Both results had high recall (sensitivity), reaching 89.7% for MMSE (macro-recall) and 91.3% for CDR (macro-recall). MMSE was more frequently mentioned in the notes and ChatGPT’s macro precision (PPV) was 82.7%. CDR, on the other hand, was less frequent, and we observed that ChatGPT hallucinates (factitiously generates) results occasionally leading to a macro precision of only 57.5%. LlaMA-2 results were significantly lower than that of ChatGPT across all metrics. A detailed qualitative analysis of the ChatGPT errors for both CDR and MMSE, and LlaMA-2 results for MMSE are included in Supplementary section S5. The majority of the errors corresponded to ChatGPT presenting results of another test instead of the one indicated as the answer. LlaMA-2 had higher rate of unexplained hallucinations. Taking positive and negative results into account, overall, ChatGPT had the highest performance with MMSE and CDR results being 83% and 89% accurate according to the double-reviewed notes.

## Discussion

In this study, our primary objective was to evaluate the performance of two state of the art LLMs (ChatGPT and LlaMA-2), in extracting information from clinical notes, specifically focusing on cognitive tests such as the Mini-Mental State Examination (MMSE) and Clinical Dementia Rating (CDR). Our results revealed that ChatGPT achieves high accuracy in extracting relevant information for MMSE and CDR scores, as well as their associated dates, with high recall, capturing nearly all of the pertinent details present in the clinical notes. The overall accuracy of ChatGPT in information extraction for MMSE and CDR were 83% and 89% respectively. The extraction was highly and had outstanding true-negative-rates. The precision of the extracted information was also high for MMSE although in the case of CDR, we observed that ChatGPT occasionally mistook other tests for CDR. Based on the ground-truth provided by our reviewers, 89.1% of the notes included an MMSE documentation instance, whereas only 14.3% of the notes included a CDR documentation instance. This, combined with our analysis of the errors, explain lower precision in the CDR case, and suggest combining ChatGPT with basic NLP preprocessing may improve the LLM performance further. Compared to ChatGPT, the open-source state of the art LLM (LlaMA-2) achieved lower performance across all metrics. The substantial inter-rater-agreement among our expert reviewers further supported the robustness and validity of our findings, and the reviewers considered ChatGPT’s responses correct and complete.

The findings of our study demonstrate that ChatGPT (powered by GPT-4), offer a promising solution for extracting valuable clinical information from unstructured notes. This approach provides a more efficient and scalable approach compared to previous methods that either rely on rigid rule-based systems or involve training resource intensive task specific models. Validated and accurate LLMs such as ChatGPT can be effortlessly applied to enhance the value of clinical data for research, enable harmonization with disease registries and biobanks, improve outreach programs within health centers, and contribute to the advancement of precision medicine. Additionally, the availability of large labeled datasets resulting from this information extraction process can also enable AI models to be trained for a wide variety of tasks.

Furthermore, our findings have implications for future AD/ADRD research. Currently, the majority of research in scalable development and validation of AI tools for early AD/ADRD detection rely on research cohorts. These cohorts are overwhelmingly white (NACC cohort is 83% white [68] ADNI cohort is 92% white [62], and do not represent true at-risk populations who tend to have higher comorbid disease burden [50]. Due to late detection and diagnosis of AD/ADRD [46] [47] [48] [49], clinical data often lacked the details necessary for accurate case identification (i.e. structured data such as ICD codes would yield low sensitivities). Using LLMs to extract data from clinical notes has the potential to improve the quality of clinical data, paving the way for clinical validation and development of clinically applicable novel AI tools and performing cognitive-health precision medicine at scale.

## Limitations

Our focus was on evaluating the information extraction capabilities of two current state of the art of LLMs, specifically ChatGPT powered by GPT-4, and LlaMA-2, rather than comparing it to all other LLMs or NLP methods. We believe that our results may be enhanced with better prompt engineering and combining LLMs with standard NLP. Additionally, we conducted a large-scale human evaluation for a single dementia use case, prioritizing result reliability over assessing various clinical scenarios. It is also important to note that our findings pertain specifically to information retrieval from clinical notes and do not predict how LLMs will perform on medical tasks requiring diagnosis, treatment recommendation, or summarization.

## Conclusions

In this diagnostic/prognostic study of ChatGPT and LlaMA-2 for extracting cognitive exam dates and scores from clinical notes, ChatGPT exhibited high accuracy in extracting MMSE scores and dates, with better performance compared to LlaMA-2. The use of LLMs could benefit dementia research and clinical care, by identifying eligible patients for treatments initialization or clinical trial enrollments. Rigorous evaluation of LLMs is crucial to understanding their capabilities and limitations.

## Data Availability

The data contains private patient information and will not be made available.

## Data Sharing Statement

The original clinical notes will not be shared, however ChatGPT and LlaMA-2 json results as well as the manually produced ground truth can be made available upon request.

## Acknowledgements

This study was supported by NYU Langone Medical Center Information Technology (MCIT) center. Author N.R. and A.M. are also supported by the following awards National Institute On Aging, of the National Institutes of Health, under Award Numbers R01AG085617 and P30AG066512. Authors H.Z. S.J., V.J.M, J.A.D., A.A.B., Y.A., A.M. and N.R. are also supported by the award number R01AG079175 from the National Institute On Aging, of the National Institutes of Health.

The following 22 authors are our clinical reviewers who also contributed to reviewing and authorship of the manuscript: N.G, I.S.J, A.B.C, N.H., N.F.A, L.J.B., A.J.C., Z.K., E.C.S., J. P., J.S., K.L., B.S., S.M., E.J.K., J.L., T.M.H, A.A., E.C., J.D., W.S., E.C.; Authors N.J., V.J.M. H.G., and Y.A., provided significant contributions to dataset construction, Redcap evaluation design and analysis and writing. Author Simon Jones performed statistical analysis. Authors J.A.D., A.A.B., and A.M. provided significant domain expertise in conceptualization and assistance in writing. Author N.R. led the study, assembled the team, and supervised the full execution of the study and is the corresponding author. Author H.Z. completed all Llama-2 analysis and helped in writing.

