## Supplementary File for "Evaluating Large Language Models in Extracting Cognitive Exam Dates and Scores"

**Section S1. Characteristics of Patients with Cognitive Tests With and Without MRI in the System**

Table S1 - Characteristics of patients with a cognitive score with or without an MRI, vs the subset of those with an MRI in the system.

| **Feature** | **Patients with cognitive score and with or without MRI in the system N=52949** | **Patients with cognitive score and an MRI, selected for the study N=458** |
| --- | --- | --- |
| **Patient demographics** | | |
| Age at time of note (mean(sd)) | 69.64 (12.5) | 73.68 (14.01) |
| Gender | | |
| Female (%) | 47.84 % | 52.54 % |
| Male(%) | 38.77 % | 47.46 % |
| Unknown or X | 13.39 % | 0 |
| Race | | |
| Asian | 4.86 % | 5.90 % |
| Black | 9.85 % | 8.52 % |
| White | 55.16 % | 72.93 % |
| American Indian | 0.77 % | 0.22 % |
| Unknown | 29.36 % | 12.45 % |

**Section S2. Two examples of the notes and corresponding output that is produced by ChatGPT. (Dates for each patient note and ChatGPT responses are shifted by a random year and month, to preserve anonymous nature of notes. All note shifts for one patients are consistent.)**

**Note Example 1:**

Mar 22, 2023: Patient was last seen in 4/2022. Since last visit, she had brain MRI which showed no acute intracranial pathology. Few SVID. Her EEG is normal. She has not done neuropsych test yet due to insurance coverage.   Today, she reports her headache and dizziness improved since last visit. Her short term memory impairment is still active, affects her daily activities. No disorientation, confusion or hallucination.    History:  Patient had Covid-19 infection in 5/2021. She  has frequent headache , dizziness, brain foggy since then. She forgot thing happened minutes ago and repeated same questions frequently. She has word finding difficulty during conversation. She has short attention span.   MMSE was 28/30 in last visit.

**ChatGPT response:**

{

"MMSE": [

   {      "score": "28/30",

           "date": "4/2022"    }

   ],

  "CDR": []

}

**Note Example 2:**

January 2016: [……] Cognitive testing was repeated:         Pre CLD Post CLD     DATE  10/09/16  09/05/16  05/10/17  09/04/17  09/07/17  08/09/17  11/30/17    GDS  2  2  2  2  2   2    MMS  30  30  29  30    30    MOCA  30  29  26     30               Written Arithmetic  1       1    Number Writing  1       1    Spelling  1       1    Clock  1       1               DSST    53  56  58  58  51    PSPC  91   58  55  56  58  64               Trail Making A             trial 1 31   32.9  35.6  32.6  29.6  38.3     trial 2    36  25.1  30.7      trial 3    27.8  21.9  30.3      trial 4    29.6  27.7       mean 31   32.9  32.3  26.8  30.2  38.3     errors   0   1  0  1               Trail Making B            trial 1 55.0   84.3  58.7  53.4  62.7  59.7     trial 2    60.2  76.2  46.7      trial 3    57  61.2  56.2      trial 4    39.9  52.5  73.3      mean 55.0   84.3  54.0  60.8  59.7  59.7     errors   1  1  3  3  0        118/78@78 bpm, RR = 18

**ChatGPT Response:**

 ChatGPT Response:

 "MMSE": [

    {

      "score": 30,

      "date": "January 2016"

    },

    {

      "score": 30,

      "date": "09/05/17"

    },

    {      "score": 29,

      "date": "05/10/18"

    },

    {

      "score": 30,

      "date": "09/04/18"

    },

    {

      "score": 30,

      "date": "09/07/18"

    },

{

      "score": 30,

      "date": "11/30/18"

 }

  ],

  "CDR": []}

**Section S3. Parsing the JSON results**

ChatGPT and LlaMA-2 were instructed to provide answers in JSON format. The human expert reviewers were instructed to provide ground truth data also in JSON format. To facilitate the review process for expert reviewers and reduce errors due to word choices, human reviewers were recommended to start from ChatGPT’s answer and correct the incorrect parts. Both results were then parsed automatically and turned into a list of (score, date) tuples, (four lists of tuples per note: 1) ChatGPT’s MMSE, 2) ChatGPT’s CDR, 3) reviewer’s MMSE, 4) reviewer’s CDR.) MMSE and CDR scores were analyzed individually. To normalize the JSON entries, any entry corresponding to the “score” which had obvious wrong test types (i.e “MoCA”, “GDS”) was automatically excluded.

**Section S4. JSON responses**

Two (out of 722) responses from ChatGPT were not in correct JSON format and failed automatic parsing. These two cases were both reporting “no MMSE or CDR.” Another 26 of the remaining 722 ChatGPT JSON results had “MMSE_Scores” and “CDR_Scores” as the JSON entry, while the rest had “MMSE” and “CDR” as the entries. The JSON results of 79 notes had additional entries, primarily including other cognitive tests (47 “MoCA” and 20 “GDS” being the most frequent wrong tests). These additional entries in JSON results of both ChatGPT and human reviewers were automatically excluded from analysis.

All LlaMA2 responses were in correct JSON format and were analyzed in the exact same manner as the responses from ChatGPT.

Of all 1031 (309×2 + 413) human expert responses, 131 were not in correct JSON format and failed automatic parsing. Most, 92, of those were missing a single formatting character (e.g. comma, bracket) and were manually corrected. The remaining 39 were long entries not provided in JSON format that would have required substantial edits and were consequently excluded from analysis. As a result, 12 of the notes (3 coming from the double-reviewed notes) were excluded from further analysis. In the remaining 710 notes (306 of which were double-reviewed), all parsed ground truth and ChatGPT and LlaMA2 instances of MMSE and CDR tests were analyzed for computing accuracy, precision, and recall. ChatGPT’s and LlaMA-2’s (micro- and macro-) precision and recall for MMSE and CDR items are included in Table 4.

**Section S5. Qualitative analysis of error instances of ChatGPT**

We analyzed 31 notes from the double-reviewed notes where ChatGPT’s predictions for CDR had any incorrect elements in them. In 3 notes, there was no sign of a CDR score and the result was hallucinated without an obvious reason. In 4 cases, the date was wrong but the score was correct. The remaining 24 notes’ errors were all due to GPT mistakenly reporting results of another test (e.g. GDS, CD4, and Cup/Disk Ratio, also abbreviated CDR) instead of CDR. We similarly analyzed the 52 notes which had any errors for the MMSE predicted list. These notes’ errors included 19 with an error only in the reported date while the score was correct; 17 cases of wrong test scores (i.e mostly MoCA, only 2 MiniCog and 1 MSK cases) being picked instead of MMSE; 4 cases of inconsistency in the note itself; 2 typos (one case where the note said “AOx3” for the score, another where MMSE was written as “MMS E”) according to the reviewer comments; 3 cases of hallucinations without specific reasons; 4 cases of non-numeric MMSE values (i.e. “deferred'' or “preserved”); 1 case of reporting partial MMSE scores instead of the total MMSE score; 1 case was missed from a table that listed 7 MMSE score/dates that were easy to see in the HTML version of the note but confusing in text-only version; and finally, 1 case was actually correct and the error was due to not considering “not found”' equal to an empty list.

For MMSE, we also reviewed the errors of LlaMA-2. Among the 98 double-reviewed notes with any error in the results according to LlaMA-2, we observed the following errors: 27 cases had total hallucination. In 19 cases other scores were reported instead of MMSE. There were 25 missed scores, and 23 cases where the wrong date was reported for the right score.

Supplementary Table S1: Diagnosis Criteria for Cognitively Normal, Mild Cognitive Impairment and Mild Alzheimer’s Disease Dementia in ADNI cohorts.

| Criteria | ADNI |
| --- | --- |
| Cogniti-vely Normal | ● No Memory Complaints aside from those common to other normal subjects of that age range.  ● Normal memory function documented by scoring at specific cutoffs on the Logical Memory II subscale (delayed Paragraph Recall) from the Wechsler Memory Scale - Revised (the maximum score is 25): a) greater than or equal to 9 for 16 or more years of education b) greater than or equal to 5 for 8-15 years of education c) greater than or equal to 3 for 0-7 years of education.  ● Mini-Mental State Exam score between 24 and 30 (inclusive) (Exceptions may be made for subjects with less than 8 years of education at the discretion of the project director).  ● Clinical Dementia Rating = 0. Memory Box score must be 0.  ● Cognitively normal, based on an absence of significant impairment in cognitive functions or activities of daily living. |
| MCI | ● Memory complaint by subject or study partner that is verified by a study partner.  ● Abnormal memory function documented by scoring below the education adjusted cutoff on the Logical Memory II subscale (Delayed Paragraph Recall) from the Wechsler Memory Scale – Revised (the maximum score is 25): a) less than or equal to 8 for 16 or more years of education b) less than or equal to 4 for 8-15 years of education c) less than or equal to 2 for 0-7 years of education.  ● Mini-Mental State Exam score between 24 and 30 (inclusive) (Exceptions may be made for subjects with less than 8 years of education at the discretion of the project director).  ● Clinical Dementia Rating = 0.5. Memory Box score must be at least 0.5.  ● General cognition and functional performance sufficiently preserved such that a diagnosis of Alzheimer’s disease cannot be made by the site physician at the time of the screening visit. |
| AD | ● Memory complaint by subject or study partner that is verified by a study partner.  ● Abnormal memory function documented by scoring below the education adjusted cutoff on the Logical Memory II subscale (Delayed Paragraph Recall) from the Wechsler Memory Scale – Revised (the maximum score is 25): a) less than or equal to 8 for 16 or more years of education b) less than or equal to 4 for 8-15 years of education c) less than or equal to 2 for 0-7 years of education.  ● MMSE between 20 and 26 (inclusive) (Exceptions may be made for subjects with less than 8 years of education at the discretion of the protocol PI).  ● Clinical Dementia Rating = 0.5, 1.0 ➪ NINCDS/ADRDA criteria for probable AD. |

Supplementary Table S2: Description of the errors encountered by ChatGPT API

| Error | Meaning |
| --- | --- |
| Azure content management violation | The content of the note activated the “content filtration” classifier that monitors content sent to the ChatGPT API for harmful content or content that violates Asure’s usage policy.  No additional information is made available to us about what aspect of the clinical note triggered this classifier.  Azure OpenAI Service includes a content filtering system that works alongside core models. This system works by running both the prompt and completion through an ensemble of classification models aimed at detecting and preventing the output of harmful content. The content filtering system detects and takes action on specific categories of potentially harmful content in both input prompts and output completions. Variations in API configurations and application design may affect completions and thus filtering behavior. |
| API timeouts | When making requests to the OpenAI API, there is a predefined time period within which the API expects to provide a response. If the API fails to respond within that time frame, it generates a "Request timed out" error.  This error may be due to reaching limits of number of requests per second, or other factors. |
| Maximum length limit error | ChatGPT 8K allows 8000 tokens in the input. This error occurs when then input includes more than 8K tokens in the input. |

Supplementary Table S3: The prompt (and the full request JSON for the task) for ChatGPT. CLINICAL_NOTE would include the date of the note (from EPIC) + “:” + the text-only content of the notes.

openai.api_type = "azure"

openai.api_version = "2023-03-15-preview"

response = openai.ChatCompletion.create(

engine="GPT4",

messages = [{"role":"system",

"**content**":"You are an AI assistant that helps doctors find information from recorded clinical notes that talk about the cognitive health of patients. \n\nPlease identify and extract all instances of Mini Mental Status Exam (MMSE) and Cognitive Dementia Rating (CDR) scores mentioned in the given text, along with the dates that you think the tests were administered, and provide the information in JSON format."}, {"**role**":"user","**content**":CLINICAL_NOTE}],

temperature=0,

max_tokens=800,

top_p=0.95,

frequency_penalty=0,

presence_penalty=0,

stop=None)

Supplementary Table S4: The prompt (and the full request JSON for the task) for LlaMA-2. CLINICAL_NOTE would include the date of the note (from EPIC) + “:” + the text-only content of the notes.

system_message = """You are an AI assistant that helps doctors find information from recorded clinical notes that talk about cognitive health of patients.

In JSON format, please identify and extract all instances of Mini Mental Status Exam (MMSE) and Cognitive Dementia Rating (CDR) scores mentioned in the given text, along with the dates that you think the tests were administrated.

If the score doesn't exist, please leave it blank.

Only include the score for MMSE or CDR if there's a numeric value, otherwise do not include the entry.

No need to attach explanations after the JSON, also don't include scores that are not MMSE or CDR.

I'm also attaching the date of when the note is written to help you better with interpreting the correct date, those will be in the beginning of the text.

The output should look something like this:

{

"MMSE": [

{

"score": "",

"date": ""

}

],

"CDR": [

{

"score": "",

"date": ""

}

]

}

"""

template = """

<s>[INST] <<SYS>>

{system_message}

<</SYS>>

{user_message} [/INST]

"""

user_message = row['note']

date = row['date']

prompt = template.format(system_message=system_message, user_message=f"{date} {user_message}")

model_input = tokenizer(prompt, return_tensors="pt").to('cuda')

LlaMA2_response = model.generate(**model_input, max_new_tokens=800, temperature=0.4, top_p=0.95, do_sample=True)
